# Sex Differences in Pulmonary Hypertension and Associated Right Ventricular Dysfunction

**DOI:** 10.1101/2024.04.25.24306398

**Authors:** Janet I. Ma, Ndidi Owunna, Nona M. Jiang, Xiaodan Huo, Emily Zern, Jenna N. McNeill, Emily S. Lau, Eugene Pomerantsev, Michael H. Picard, Dongyu Wang, Jennifer E. Ho

## Abstract

**Background:** Prior studies have established the impact of sex differences on pulmonary arterial hypertension (PAH). However, it remains unclear whether these sex differences extend to other hemodynamic subtypes of pulmonary hypertension (PH).

**Methods:** We examined sex differences in PH and hemodynamic PH subtypes in a hospital-based cohort of individuals who underwent right heart catheterization between 2005-2016. We utilized multivariable linear regression to assess the association of sex with hemodynamic indices of RV function [PA pulsatility index (PAPi), RV stroke work index (RVSWI), and right atrial: pulmonary capillary wedge pressure ratio (RA:PCWP)]. We then used Cox regression models to examine the association between sex and clinical outcomes among those with PH.

**Results:** Among 5208 individuals with PH (mean age 64 years, 39% women), there was no significant sex difference in prevalence of PH overall. However, when stratified by PH subtype, 31% of women vs 22% of men had pre-capillary (P<0.001), 39% vs 51% had post-capillary (P=0.03), and 30% vs 27% had mixed PH (P=0.08). Female sex was associated with better RV function by hemodynamic indices, including higher PAPi and RVSWI, and lower RA:PCWP ratio (P<0.001 for all). Over 7.3 years of follow-up, female sex was associated with a lower risk of heart failure hospitalization (HR 0.83, CI 95% CI 0.74– 0.91, p value <0.001).

**Conclusions:** Across a broad hospital-based sample, more women had pre-capillary and more men had post-capillary PH. Compared with men, women with PH had better hemodynamic indices of RV function and a lower risk of HF hospitalization.

**CLINICAL PERSPECTIVE:**

1. What Is New?

- Although sex differences have been explored in pulmonary arterial hypertension, sex differences across pulmonary hypertension (PH) in broader samples inclusive of all hemodynamic subtypes remain less well defined
- We delineate sex differences in hemodynamic subtypes of PH and associated right ventricular function in a large, heterogenous, hospital-based sample of individuals who underwent right heart catheterization
- Sex has a significant impact on prevalence of PH across hemodynamic subtypes as well as associated RV function
2. What Are the Clinical Implications?

- Understanding sex differences across different PH hemodynamic subtypes is paramount to refining risk stratification between men and women
- Further elucidating sex differences in associated RV function and clinical outcomes may aid in developing sex-specific therapies or management strategies to improve clinical outcomes

## INTRODUCTION

Pulmonary hypertension (PH) is a heterogenous disease associated with poor clinical outcomes (1). Clinically, PH can be classified by World Health Organization groups based on etiology. PH is also often grouped by hemodynamic assessment into precapillary PH, postcapillary PH, and mixed PH, which carry significant prognostic and therapeutic implications, particularly for PH with a precapillary component (1). In addition, one of the strongest predictors of mortality in this patient population is RV dysfunction, a frequent long-term consequence of PH and associated with poor prognosis (2).

Sex differences have long been observed in pulmonary arterial hypertension (PAH), a subset of PH also known as WHO Group 1 PH with precapillary hemodynamic characteristics. Specifically, PAH is noted to have a much higher prevalence in women compared with men, and yet men appear to have worse RV dysfunction and worse survival, suggesting sex-specific phenotypes with regards to associated RV dysfunction (3). The etiology of these differences remains unclear, but some have postulated that sex hormones such as higher levels of estrogen may be responsible for both the increased risk women face in developing PAH as well as their improved RV function and survival compared with men (3).

Despite the extensive study of sex differences in prevalence and clinical trajectory of PAH, little is still known about sex differences across other hemodynamic PH subtypes. In this context, we sought to study sex differences in PH prevalence, associated RV dysfunction, and clinical outcomes across a broad and clinically heterogeneous hospital-based sample of individuals undergoing right heart catheterization (RHC). Based on what is known about PAH, we hypothesized greater prevalence in women compared with men in precapillary but not postcapillary PH, as well as better RV function and clinical outcomes in women compared with men across all hemodynamic PH subtypes.

## METHODS

### Study Sample

Our study examined ambulatory and hospitalized patients who underwent right heart catheterization (RHC) between 2005 and 2016 at Massachusetts General Hospital. If an individual underwent multiple RHC procedures during this time period, only the initial RHC was included for analysis. A total of 10,306 cases were included. Cases were excluded if they met the following clinical exclusion criteria: acute myocardial infarction (MI) occurring on the same day as catheterization, cardiac arrest or shock within 24 hours, presence of mechanical ventilation, presence of intra-aortic balloon pump, history of heart or lung transplant, complicated adult congenital heart disease, history of valvular replacement, or those on dialysis (n=887). Cases were also excluded if there were missing key clinical covariates (n=484), patient identifier variables (n=398), or hemodynamic parameters (n=252), which led to a final study sample of 8285 cases. This study was approved by the appropriate Institutional Review Board.

### Clinical, PH, and RV hemodynamic variables

The following clinical characteristics were extracted from the medical records at the time of RHC: age, sex, body mass index (BMI), smoking status, and presence of comorbidities (diabetes mellitus, hypertension, history of MI, history of heart failure, prior lung disease, and chronic kidney disease). OSA was ascertained from the electronic medical record utilizing appropriate *International Classification of Diseases Ninth Revision* (*ICD-9*) or *Tenth Revision* (*ICD-10*) codes.

Hemodynamic measures were recorded directly from the time of RHC, including resting blood pressure, heart rate, mean right atrial (RA) pressure, pulmonary artery (PA) systolic and diastolic pressure, trans-pulmonary gradient (TPG), and mean pulmonary capillary wedge pressure (PCWP). Any nonphysiologic parameters identified in our database were set to missing. The PA pulsatility index (PAPi) was calculated as 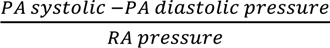. If RA pressure was recorded as zero, the value was set to one to allow for PAPi calculation. If RA pressure was recorded as negative, they were deemed non-physiologic and set to missing, which affected <1% of measurements. Cardiac output and index measurements were obtained via thermodilution methods whenever possible. If not available, assumed Fick cardiac output and index were used. Both were indexed using the Mosteller formula for body surface area [7]. Right ventricular stroke work index (RVSWI) was derived as *0.0136 x stroke volume index x (Mean PA pressure -mean RA pressure)*, where stroke volume index was calculated by dividing cardiac index by heart rate.

Pulmonary hypertension (PH) was defined as mean pulmonary arterial pressure (mPAP) >20 mmHg based on the 2022 European Society of Cardiology/European Respiratory Society guidelines (1). We further categorized into three hemodynamic subtypes: precapillary PH (TPG >12 mmHg and PCWP ≤ 15 mmHg), postcapillary PH (TPG ≤ 12 mmHg and PCWP > 15 mmHg), and mixed PH (TPG > 12 mmHg and PCWP > 15 mmHg).We utilized TPG cutpoints rather than pulmonary vascular resistance (PVR), as not all participants had available cardiac output,. For those with cardiac output data, we calculated pulmonary vascular resistance (PVR) and examined PH hemodynamic subtypes using accepted PVR instead of TPG cutpoints in secondary analyses.

### Clinical Outcomes

Primary clinical outcomes included all-cause mortality and heart failure hospitalization. All-cause mortality was ascertained using the National Social Security Death Master Index and hospital records, abstracted on 06/10/2020. Due to confidentiality purposes, the precise dates of deaths that occurred between 06/10/2017 and the date of abstraction are protected nationally. Time-to-event analyses were conducted by imputing death dates at the midpoint of the blanking period (12/10/2018). Heart failure hospitalizations were defined using an ICD-9 or ICD-10 code for heart failure as the primary discharge diagnosis or a current procedural terminology (CPT) code for heart transplantation (OHT) or durable ventricular assist device (VAD). The follow-up period for each participant was defined as time from RHC to death date or date of final encounter in the electronic health record. Patients were censored based on time of last encounter.

### Statistical Analysis

Baseline characteristics were summarized across the total sample stratified by sex. Sex differences were examined in prevalence of PH and PH subtypes. We then examined cross-sectional associations of sex with hemodynamic indices of RV dysfunction among those with PH, including PAPi, RA:PCWP ratio, and RVSWI using multivariable linear regression. Models were adjusted for the following clinical covariates: age, sex, BMI, hypertension, diabetes mellitus, obstructive sleep apnea (OSA), chronic lung disease, prevalent MI, prevalent HF, chronic kidney disease.

We next examined the association of sex with clinical outcomes among individuals with PH using the Kaplan-Meier method. We constructed multivariable Cox models to examine the association of sex with outcomes, adjusting for age, BMI, previous MI, previous HF, hypertension, diabetes mellitus, obstructive sleep apnea (OSA), chronic lung disease, and chronic kidney disease. In secondary models, we stratified by PH hemodynamic subtypes. In exploratory analyses, we evaluated whether sex modified the association of RV function with adverse outcomes among those with PH using multiplicative interaction terms (sex*PAPi, sex*RVSWI, and sex*RA:PCWP ratio). Analyses were conducted using SAS software, Version 9.4 (Cary, North Carolina, USA).

## RESULTS

### Sex differences in prevalence of PH and hemodynamic subtypes

Among 8285 individuals who underwent right heart catheterization between 2005-2016, 5208 (63%) met criteria for PH, as defined by a mean pulmonary arterial pressure (mPAP) >20 mmHg. There was no significant sex difference in the prevalence of overall PH, with n= 2032 (62%) women and n=3176 (63%) men meeting PH criteria (p=0.96, baseline characteristics in **Supplemental Table 1**).

Among individuals with PH, average age was 64±12 years and 39% were women. Men were significantly more likely to have comorbid conditions including previous MI, previous HF, hypertension, diabetes mellitus, hypercholesteremia, and OSA (p <0.001 for all). However, women were more likely to have chronic lung disease compared with men (20.2% vs 16.7%, p=0.001). The remaining baseline characteristics of individuals with PH are shown in **Table 1**.

**Table 1.**
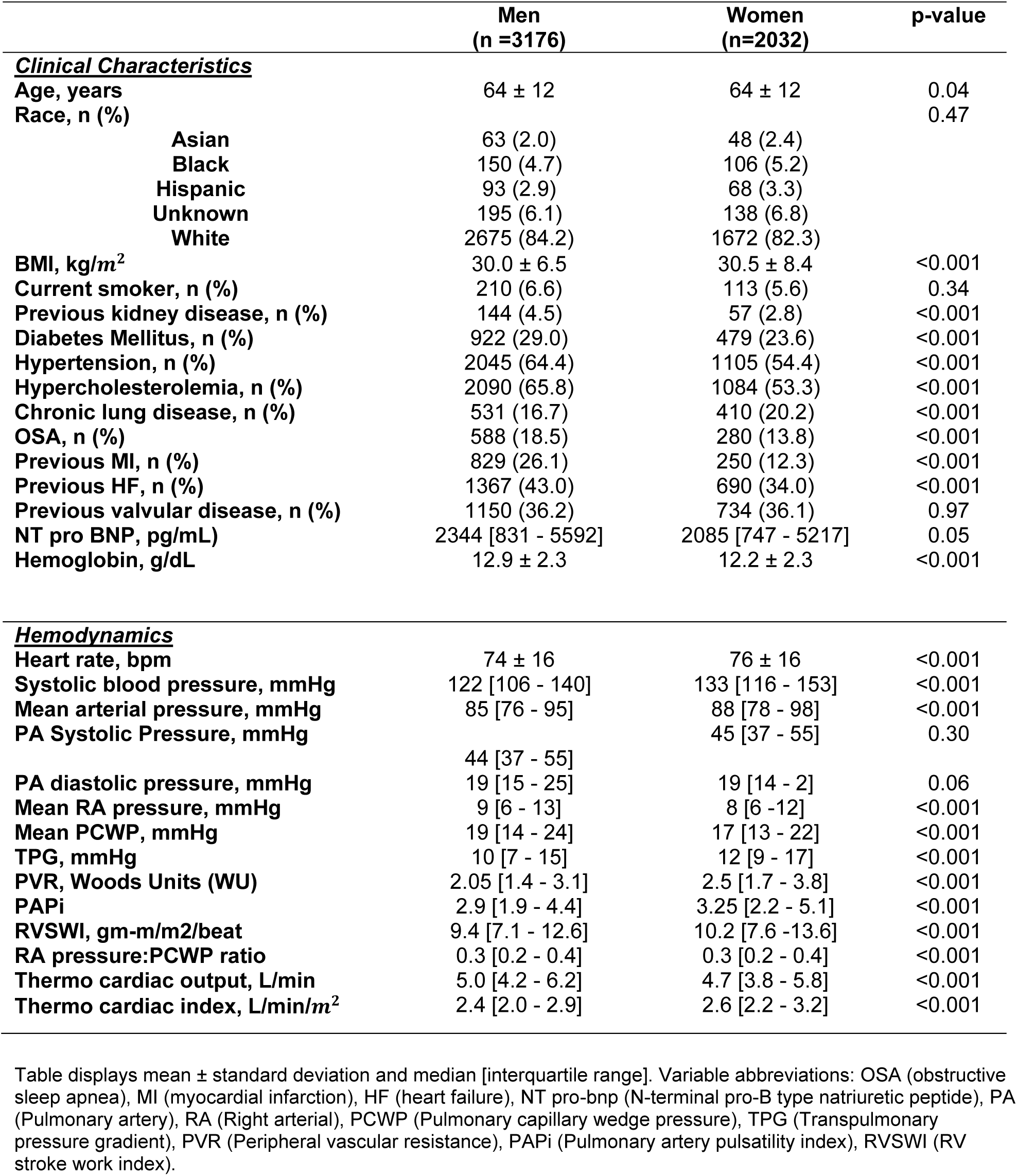
Clinical characteristics of men and women with PH, n= 5208.

We found that hemodynamic PH subtypes differed significantly by sex (**Figure 1**). Overall, women were more likely to have pre-capillary PH and men were more likely to have post-capillary PH. Specifically, 26% of women vs 19% of men had pre-capillary (P<0.001), 34% vs 44% had post-capillary (P 0.03), and 26% vs 23% had mixed PH (P=0.08).

**Figure 1.**
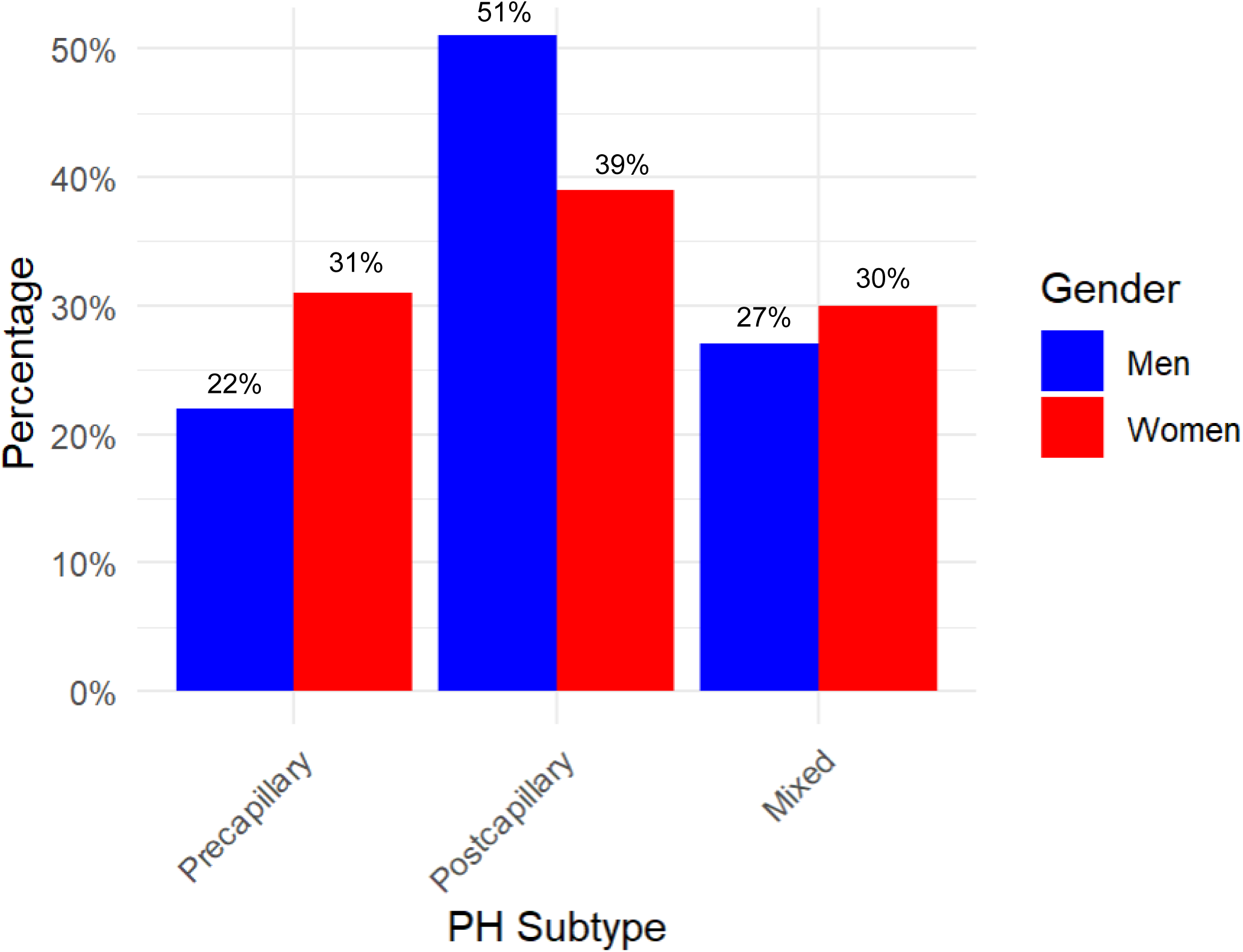
Hemodynamic PH subtypes among women and men with PH. Prevalence of hemodynamic PH subtypes in women and men. Among women with PH, 31% (n = 544) had precapillary PH, 39% had post-capillary PH (n = 685), and 30% had mixed (n = 527). Among women with PH, 22% (n = 605) had precapillary PH, 51% had post-capillary PH (n = 1402), and 27% had mixed (n = 743). Note that total individuals with precapillary, postcapillary, and mixed PH in our study was 4506 out of 5208 total individuals with PH; the remaining 702 individuals with PH who did not meet criteria for one of these hemodynamic PH subtypes were excluded in this figure.

### Sex differences in RV function among individuals with PH

Among individuals with PH, female sex was associated with better RV function as ascertained using hemodynamic indices (**Table 2**). Specifically, women with PH had higher PAPi (β 0.18, SE −0.03, p <0.001), lower RA:PCWP ratio (β −0.18, SE −0.03, p <0.001), and higher RVSWI (β 0.17, SE −0.03, p <0.001) in multivariable adjusted analyses.

**Table 2.** Association of female sex with indices of RV Function in individuals with PH.

| **Sample** | **Measures of RV Function** | **β-estimate** | **Standard Error** | **p-value** |
| --- | --- | --- | --- | --- |
| **Overall PH** | **PAPi** | 0.18 | -0.03 | <0.001 |
|  | **RA:PCWP ratio** | -0.18 | -0.03 | <0.001 |
|  | **RVSWI** | 0.17 | -0.03 | <0.001 |
| **Pre-capillary PH** | **PAPi** | 0.13 | 0.03 | <0.001 |
|  | **RA:PCWP ratio** | -0.17 | 0.03 | <0.001 |
|  | **RVSWI** | 0.01 | 0.03 | 0.74 |
| **Mixed PH** | **PAPi** | 0.12 | 0.03 | <0.001 |
|  | **RA:PCWP ratio** | -0.15 | 0.03 | <0.001 |
|  | **RVSWI** | 0.03 | 0.03 | 0.33 |
| **Post-capillary PH** | **PAPi** | 0.10 | 0.03 | <0.001 |
|  | **RA:PCWP ratio** | -0.14 | 0.03 | <0.001 |
|  | **RVSWI** | 0.04 | 0.03 | 0.13 |

$\beta$ -Estimate represents standard deviation unit difference in RV outcome measure between women vs men. Multivariable adjusted model was adjusted for age, BMI, prior heart failure, hypertension, diabetes, OSA, Chronic Lung Disease, previous MI, prior kidney failure, and chronic heart failure history. Variable abbreviations: RA: PCWP Ratio (Right Arterial/Pulmonary capillary wedge pressure ratio), PAPi (pulmonary artery pulsatility index), RVSWI (RV stroke work index); PH, pulmonary hypertension.

When stratified across PH hemodynamic subtypes, these sex differences in RV function persisted for PAPi and RA:PCWP but not for RVSWi (**Table 2**). Women had higher PAPi compared with men in pre-capillary PH (β 0.13, SE 0.03, p <0.001), post-capillary PH (β 0.1, SE 0.03, p <0.001), and mixed PH (β 0.12, SE 0.03, p <0.001) (**Supplemental figure 1**). Women also had lower RA:PCWP ratio in pre-capillary PH (β −0.17, SE 0.03, p <0.001), post-capillary PH (β −0.18, SE 0.03, p <0.001), and mixed PH (β −0.15, SE 0.03, p <0.001).

### Sex differences in clinical outcomes for PH

We next examined the association of sex with clinical outcomes among individuals with PH. Over a median follow-up time of 7.3 years (IQR 4.2 – 10 years), 4023 individuals experienced HF hospitalizations (1625 among women, 2398 in men), with 3549 all-cause death events (1413 in women, 2136 in men).

In multivariable-adjusted analyses, we found that female sex was associated with 17% lower hazard of heart failure hospitalization (HR 0.83, 95% CI 0.74– 0.91, p <0.001). This was driven predominantly by individuals with postcapillary PH, where women with postcapillary PH were significantly less likely to experience HF hospitalization than men (HR 0.79, 95% CI 0.68-0.93, p-value = 0.007, **Table 3**), whereas no differences in HF hospitalization were observed among those with pre-capillary or mixed PH. Further, we observed no sex differences in risk of mortality among PH overall (HR 0.92, 95% CI 0.84-1.00, p = 0.05, **Figure 2**), nor by PH subtype.

**Figure 2.**
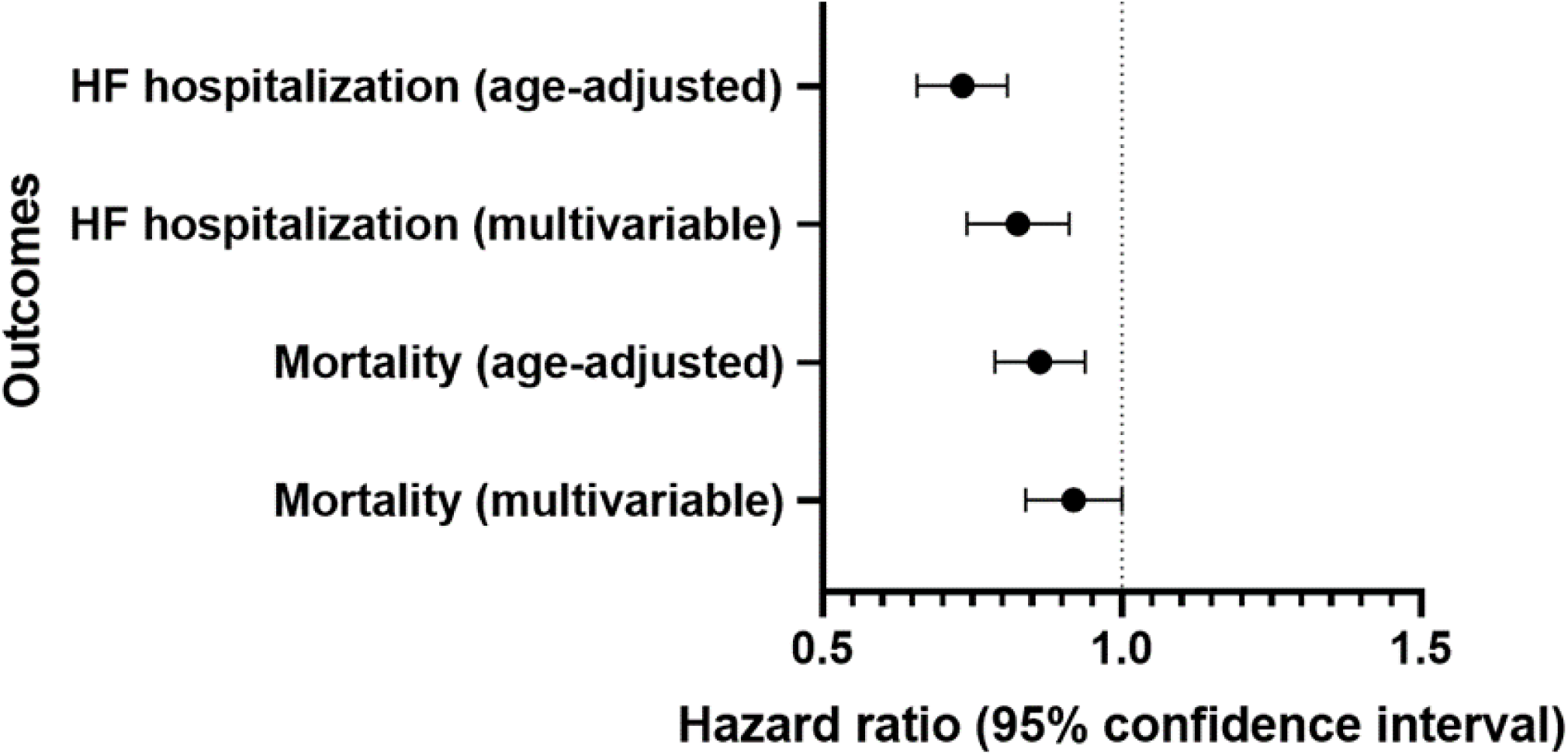
Association of female sex with adverse outcomes among individuals with PH. A forest plot demonstrating age-adjusted and multivariable analyses with confidence intervals for the association between sex and adverse outcomes, specifically heart failure hospitalization and mortality. Multivariable models were adjusted for age, sex, hypertension, diabetes mellitus, obstructive sleep apnea, chronic lung disease, prevalent myocardial infarction, prevalent heart failure.

**Table 3.** Association of female sex with clinical outcomes in individuals with PH.

| Sample | Outcome | HR | 95% CI | p-value |
| --- | --- | --- | --- | --- |
| Overall PH | HF Hospitalization | 0.83 | 0.74– 0.91 | <0.001 |
|  | All-Cause Mortality | 0.92 | 0.84 –1.00 | 0.05 |
| Pre-capillary PH | HF Hospitalization | 0.93 | 0.70-1.21 | 0.61 |
|  | All-Cause Mortality | 0.90 | 0.75-1.08 | 0.25 |
| Post-capillary PH | HF Hospitalization | 0.79 | 0.68-0.93 | 0.007 |
|  | All-Cause Mortality | 0.88 | 0.76-1.03 | 0.11 |
| Mixed PH | HF Hospitalization | 0.83 | 0.69-1.01 | 0.058 |
|  | All-Cause Mortality | 0.88 | 0.75-1.03 | 0.10 |

Hazard ratios (HR) represent comparison of adverse PH outcomes in women when compared to men. Multivariable models were adjusted for age, sex, hypertension, diabetes mellitus, obstructive sleep apnea, chronic lung disease, prevalent myocardial infarction, prevalent HF. Abbreviations: CI, confidence interval; HR, hazard ratio; HF, heart failure; PH, pulmonary hypertension

## DISCUSSION

We examined sex differences in prevalence of PH, hemodynamic PH subtypes, and associated RV dysfunction and clinical outcomes in a large hospital-based sample representing a broad spectrum of cardiopulmonary disease referred for RHC. While prevalence of overall PH was similar among women and men, women were more likely to have precapillary PH while men were more likely to have postcapillary PH. Among individuals with PH, female sex was consistently associated with better hemodynamic indices of RV function and lower risk of heart failure hospitalization. These findings build on a current body of literature that has thus far primarily explored sex differences in PAH, augmenting our understanding to a much broader population encompassing all PH hemodynamic subtypes with potential implications for future sex-specific risk stratification and treatment in these groups.

Our findings regarding prevalence extend upon well-described sex differences in PAH, a subset of precapillary PH, where female sex has been established as a primary risk factor (3, 4). While prevalence of PAH is higher among women vs men, women with PAH demonstrate better RV function and overall clinical outcomes compared with men. A systematic review of 11 randomized trials found that women with PAH had lower right atrial pressures, higher cardiac index, and lower pulmonary vascular resistance compared with men (5). Similarly, a study by Kjellstrom et al examining sex differences in individuals with idiopathic PAH found that women were younger, had lower PVR, and fewer comorbidities than men (6). They found that although women were more prevalent in PAH, men had worse survival rates although this difference was attenuated by age. Our findings somewhat differ in clinical outcomes when examining a broader group of individuals with PH across hemodynamic subtypes, where risk of heart failure hospitalization but not mortality was significantly higher for men compared with women in all-comers with PH.

Conversely, our findings regarding the predominant male prevalence in post-capillary pulmonary hypertension is noteworthy. While our study is not able to attribute exact etiology, this is likely in the setting of left-sided heart disease, where known sex differences also exist. For example, prior studies have demonstrated that men are more likely to develop HF with reduced ejection fraction (HFrEF), while women are more predisposed to HF with preserved ejection fraction (HFpEF) (7, 8). While both HF subtypes have similarly wide ranges in estimated PH prevalence (40-75% for HFrEF, and 36-83% for HFpEF, (9–14) one study found that among individuals with HFpEF, 82% who develop PH are women, compared with 58% without (12). Our finding in prevalence of postcapillary PH is in keeping with the sex differences found in baseline prevalent HF amongst all individuals with PH, a co-morbidity present in 43% of men with PH but only 34% of women with PH (p-value < 0.001).

In prior studies of sex differences in RV function, women appear to have better RV systolic function compared with men both across community-based samples without PH and among those with PAH (15, 16). For example, in the Multi-Ethnic Study of Atherosclerosis, a large prospective study examining both men and women without clinical cardiovascular disease who underwent cardiac MRI, men had higher RV mass and volumes while women had higher RV function (15).

In PAH, RV function have also been demonstrated to be higher in women compared with men. In a small retrospective cohort study of 101 individuals with idiopathic PAH by Jacobs et al, sex differences in RV function were correlated with subsequent sex differences in response to treatment and survival (17). Although RV ejection fraction was found to be similar in both men and women at baseline, it declined in men and increased in women despite comparable reductions in pulmonary vascular resistance following initiation of treatment. Men also had significantly worse transplant-free survival compared with women; a mediation analysis found 39% of this survival difference was mediated through changes in RVEF for both women and men after initiating PAH medical therapies after adjusting for potential confounders.

Other types of PH have not been studied as extensively with respect to sex differences in RV function, although there is increasing interest in elucidating distinct phenotypes between men and women. In a retrospective study by Prins et al, male sex was the strongest predictor of RV dysfunction in group 3 PH, even after adjusting for pulmonary vascular resistance (18). Men with group 3 PH also showed a progressive decline in RV contractility with increasing pulmonary vascular resistance. Similarly, among individuals with heart failure with reduced ejection fraction (LVEF <35%), men were found nearly twice as likely have concomitant RV dysfunction compared with women despite similar LV ejection fraction (19). Among individuals with heart failure with preserved ejection fraction (HFpEF), men were found to have worse RV function compared with women at every mean pulmonary arterial pressure, with male sex again being an independent predictor of RV dysfunction (20). Our findings are in keeping with these prior studies, confirming a significant sex difference in RV dysfunction across all PH hemodynamic subtypes, with women demonstrating higher RV function compared with men in precapillary, postcapillary, and mixed PH.

The mechanisms underlying sex differences in RV dysfunction and PH remain unclear, although sex hormones are postulated to play an important role since cardiac myocytes express receptors for all major sex hormones (21). The “estrogen hypothesis” that higher estrogen levels confers a protective effect in pulmonary arterial pulmonary hypertension has been touted as a possible mechanism. This has been tested across various preclinical models: in a rat model of PH, estrogen E2 appears to promote adverse pulmonary vascular remodeling but also demonstrate RV protective effects in vivo through decreased RV apoptopic signaling, pro-inflammatory cytokine expression, oxidative stress, and mitochondrial dysfunction (22–25). Other studies have demonstrated a direct role of E2 on pulmonary vasculature, particularly increasing both proximal and distal pulmonary arterial compliance, which could have a downstream effect on RV remodeling and function (26–28). In humans, estrogen appears to favorably modulate RV function. In the MESA-Right Ventricle study which examined the role of sex hormones in individuals without clinical cardiovascular disease, Ventetuolo et al showed that post-menopausal women taking estradiol (an estrogen-based steroid hormone) for menopausal hormone therapy, had higher RV ejection fraction compared with men (29).

Our study has a number of limitations worth noting. First, RHC allowed for diagnosis and hemodynamic subtyping of PH but not clinical categorization by WHO classification, and definitive etiology of PH was not possible to ascertain comprehensively. Second, we examined individuals undergoing clinically indicated RHC, which may have resulted in selection bias, and generalizability of our study findings needs to be interpreted in that context. Additionally, due to the observational study design, we acknowledge the possibility of residual confounding, and causal inferences regarding sex differences cannot be drawn.

Our study also exhibits several strengths, most notably the large sample size and detailed hemodynamic data from right heart catheterizations in a broad population across all PH hemodynamic subtypes. One unique strength of our study is our use of hemodynamic indices of RV function rather than non-invasive assessment using echocardiography or MRI compared with prior studies. PAPi, RA:PCWP ratio, and RVSWI have all been correlated with clinical outcomes (30–34). Previous literature has also demonstrated PAPi may correlate more significantly to severe clinical RV failure in post-LVAD patients compared with echocardiographic-derived measurements, although this has not been as well-established in other clinical situations (35).

In sum, we found similar prevalence of overall PH among women and men, with women more likely to have precapillary PH while men were more likely to have postcapillary PH. Further, among individuals with PH, female sex was associated with better hemodynamic indices of RV function and lower risk of heart failure hospitalization.

These findings extend prior studies predominantly focused on sex differences in PAH to a large and clinically heterogeneous hospital-based sample with invasive hemodynamic assessment. Our findings illustrate the growing need for future studies to better understand underlying mechanisms and sex-specific considerations in RV dysfunction and associated clinical outcomes for individuals across all PH subtypes, as they may have important implications for risk stratification and management.

## Data Availability

The data and study materials will not be made available to other researchers for purposes of reproducing the results or replicating the procedure. Statistical code and analytic methods will be made available upon request.

## Acknowledgments

None

---

**Supplemental Table 1.** Baseline characteristics by sex in overall population, (n= 8285)

|  | Male<br>(n=5022) | Female<br>(n=3263) | p-value |
| --- | --- | --- | --- |
| **<u>Clinical Characteristics</u>** |  |  |  |
| Age, years | 63 ± 12 | 63 ± 14 | 0.02 |
| Race, n (%) |  |  | 0.30 |
| Asian | 89 (1.7) | 75 (2.3) |  |
| Black | 184 (3.7) | 128 (3.9) |  |
| Hispanic | 154 (3.1) | 108 (3.3) |  |
| Unknown | 304 (6.1) | 214 (6.6) |  |
| White | 4291 (85.4) | 2738 (83.9) |  |
| BMI, kg/m<sup>2</sup> | 29.3 ± 6.1 | 29.3 ± 7.8 | <0.001 |
| Current smoker, n (%) | 304 (6.5) | 156 (3.3) | 0.03 |
| Previous kidney disease, n (%) | 176 (2.1) | 65 (0.8) | <0.001 |
| Diabetes Mellitus, n (%) | 1282 (15.5) | 641 (7.7) | <0.001 |
| Hypertension, n (%) | 3196 (38.6) | 1667 (20.2) | <0.001 |
| Hypercholesterolemia, n (%) | 3306 (50.0) | 1671 (25.3) | <0.001 |
| Chronic lung disease, n (%) | 763 (9.2) | 586 (7.1) | <0.001 |
| OSA, n (%) | 803 (9.7) | 351 (4.2) | <0.001 |
| Previous MI, n (%) | 1184 (14.3) | 367 (4.4) | <0.001 |
| Previous HF, n (%) | 1768 (21.3) | 888 (10.7) | <0.001 |
| Previous valvular disease, n (%) | 1673 (20.2) | 1036 (12.5) | 0.14 |
| NT pro BNP, pg/mL | 1818 [573 - 4711] | 1622 [459 - 4306] | 0.03 |
| Hemoglobin, g/dL | 13.4 ± 2.2 | 12.5 ± 2.0 | <0.001 |
| **<u>Hemodynamics</u>** |  |  |  |
| Heart rate, bpm | 72 ± 15 | 74 ± 15 | <0.001 |
| Systolic blood pressure, mmHg | 121 [106 - 137] | 131. [114 - 151] | <0.001 |
| Mean arterial pressure, mmHg | 84 [75 - 93] | 86 [77 - 96] | <0.001 |
| PA Systolic Pressure, mmHg | 36 [29 - 48] | 36 [28 - 48] | 0.22 |
| PA diastolic pressure, mmHg | 15 [10 - 21] | 14 [9 - 20] | 0.02 |
| Mean RA pressure, mmHg | 7 [4 - 11] | 6 [4 - 10] | <0.001 |
| Mean PCWP, mmHg | 14 [10 - 21] | 13 [9 - 19] | <0.001 |
| TPG, mmHg | 9 [6 - 12] | 9 [6 - 14] | <0.001 |
| PVR, Woods Units (WU) | 1.6 [1.1 - 2.5] | 2.0 [1.4 - 3.1] | <0.001 |
| PAPi | 3.3 [2.1 - 5.2] | 3.6 [2.3 - 6.1] | <0.001 |
| RVSWI, gm-m/m2/beat | 8.0 [5.9 - 10.9] | 8.3 [6.0 - 11.7] | <0.001 |
| RA pressure:PCWP ratio | 0.3 [0.2 - 0.4] | 0.3 [0.2 - 0.4] | <0.001 |
| Thermo cardiac output, L/min | 5.2 [4.3 - 6.3] | 4.8 [3.9 - 5.8] | <0.001 |
| Thermo cardiac index, L/min/m<sup>2</sup> | 2.5 [2.1 - 3.0] | 2.7 [2.2 - 3.2] | <0.001 |

Table displays mean ± standard deviation and median [interquartile range]. Variable abbreviations: OSA (obstructive sleep apnea), MI (myocardial infarction), HF (heart failure), NT pro-bnp (N-terminal pro-B type natriuretic peptide), PA (Pulmonary artery), RA (Right arterial), PCWP (Pulmonary capillary wedge pressure), TPG (Transpulmonary pressure gradient), PVR (Peripheral vascular resistance), PAPi (Pulmonary artery pulsatility index), RVSWI (RV stroke work index).

**Supplemental figure 1.**
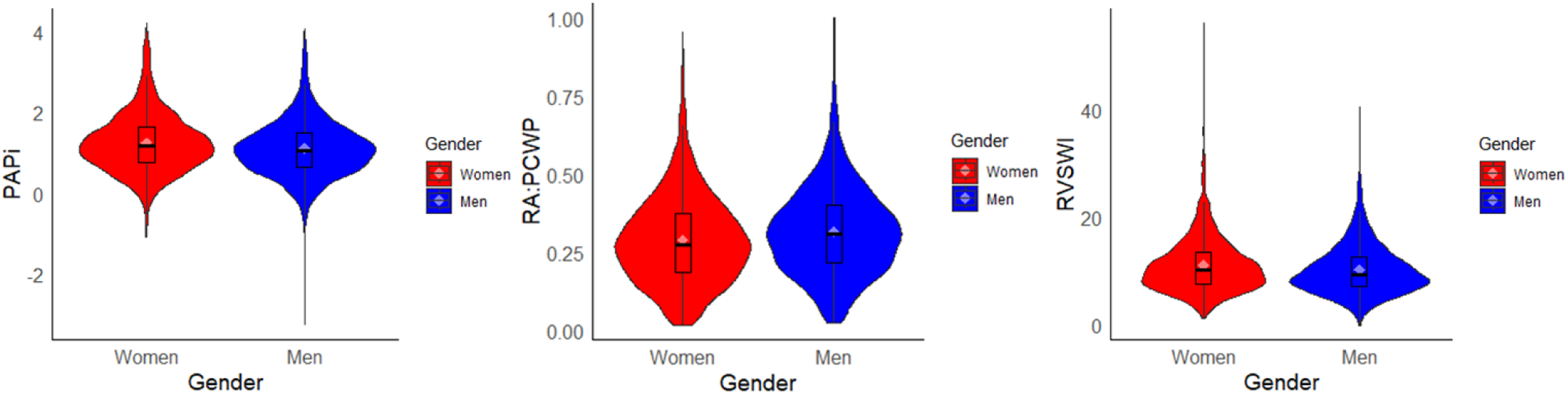
Sex differences in RV function by hemodynamic indices between women and men with PH. Violin plots demonstrating differences in RV function by hemodynamic indices (PAPi, RA:PCWP ratio, and RVSWI) stratified by men (blue) and women (red) for all individuals with PH. Variables were natural log-transformed. Women with PH had better RV function compared with men by all three indices, with higher PAPi (β 0.18, SE −0.03, p <0.001), lower RA:PCWP ratio (β −0.20, SE −0.03, p <0.001), and higher RVSWI (β 0.17, SE −0.03, p <0.001).

